# Sex-based differences in clearance of chronic *Plasmodium falciparum* infection

**DOI:** 10.1101/2020.06.10.20127720

**Authors:** Jessica Briggs, Noam Teyssier, Joaniter I. Nankabirwa, John Rek, Prasanna Jagannathan, Emmanuel Arinatiwe, Teun Bousema, Chris Drakeley, Margaret Murray, Emily Crawford, Nicholas Hathaway, Sarah G. Staedke, David Smith, Phillip J. Rosenthal, Moses Kamya, Grant Dorsey, Isabel Rodriguez-Barraquer, Bryan Greenhouse

## Abstract

Multiple studies have reported a male bias in incidence and/or prevalence of malaria infection in males compared to females. To test the hypothesis that sex-based differences in host-parasite interactions affect the epidemiology of malaria, we intensively followed *Plasmodium falciparum* infections in a cohort in a malaria endemic area of eastern Uganda and estimated both force of infection (FOI) and rate of clearance using amplicon deep-sequencing. We found no evidence of differences in behavioral risk factors, incidence of malaria, or FOI by sex. In contrast, females cleared asymptomatic infections at a faster rate than males (hazard ratio [HR] = 1.82, 95% CI 1.20 to 2.75 by clone and HR = 2.07, 95% CI 1.24 to 3.47 by infection event) in multivariate models adjusted for age, timing of infection onset, and parasite density. These findings implicate biological sex-based differences as an important factor in the host response to this globally important pathogen.

## Introduction

Malaria, a protozoan infection of the red blood cells, remains one of the greatest global health challenges^1^. Infection with malaria parasites results in a wide range of clinical disease presentations, from severe to uncomplicated; in addition, in hyperendemic areas, asymptomatic infections are common^2^. It is well established that chronic asymptomatic infection with *Plasmodium falciparum*, the most common and fatal malaria parasite, can lead to morbidity for those infected and contribute to ongoing transmission^2-5^. Characterization of these asymptomatic infections is paramount as they represent a major obstacle for malaria elimination efforts. Therefore, an understanding of how host immunity and parasite factors interact to cause disease tolerance is required. While age-specific immunity to malaria in hyperendemic areas is well-characterized, less attention has been paid to the possibility of a sex bias in malarial susceptibility despite evidence for a male bias in malaria infections in non-human animals and a male bias in the prevalence of other human parasitic infections^6-8^.

The clearest evidence for sexual dimorphism in malaria susceptibility is in pregnant women, who are at greater risk of malaria infection and also experience more severe disease and higher mortality^9^. However, multiple studies performed in different contexts have demonstrated a male bias in incidence and/or prevalence of malaria infection in school aged children and adults;^10-14^ this bias is more well established in hypoendemic areas, but has also been observed in hyperendemic regions.^15,16^ It has often been postulated that these differences in malaria incidence or prevalence stem from an increased risk of males acquiring infection due to socio-behavioral factors^11,13,17,18^. However, biological sex itself has been demonstrated to affect responses to other pathogens, and thus an alternative hypothesis is that the sexes have different responses to the malaria parasite once infected^19-22^.

Estimating the host response to *P. falciparum* infection requires close follow up of infected individuals, sensitive detection of parasites, and the ability to distinguish superinfection, which is common in endemic areas, from persistent infection. To test the hypothesis that sex-based differences in host-parasite interactions affect the epidemiology of malaria, we intensively followed a representative cohort of individuals living in a malaria endemic area of eastern Uganda. Using frequent sampling, ultrasensitive quantitative PCR (qPCR), and amplicon deep sequencing to genotype parasite clones, we were able to accurately detect the onset of new infections and follow all infections over time to estimate their duration. Using these data, we show that females cleared their asymptomatic infections more rapidly than males, implicating biological sex-based differences as important in the host response to this globally important pathogen.

## Results

### Cohort participants and *P. falciparum* infections

This analysis involved data from 477 children and adults (233 males and 244 females) that were followed for a total of 669.6 person-years (Table 1). 25 participants, 10 males and 15 females, were enrolled after initial enrollment (Figure 1); median duration of follow up in those who were dynamically enrolled was 0.84 years (IQR 0.69-1.23) compared to 1.45 years (IQR 1.43-1.47) in those enrolled during initial enrollment. 149 of 477 participants (31.2%) included in the analysis had at least one *P. falciparum* infection detected (Figure 1). 114 participants had 822 successfully genotyped samples and had infections characterized by clone and by infection event. We achieved a read count of >10,000 for 92% of genotyped samples, identifying 45 unique AMA-1 clones in our population (frequencies and sequences in **Supplemental File 1e**). 35 samples had very low-density infections (< 1 parasite/µL) that could not be genotyped and had infections characterized at the event level only. There were more baseline infections in males at the clone level, with 117/185 (63.2%) baseline infections in males and 68/185 (36.8%) baseline infections in females (p <0.001). At the infection event level, there was no difference in the number of baseline infections by sex, with 54/99 (54.4%) baseline infections in males and 45/99 (45.5%) baseline infections in females and (p = 0.26).

**Table 1.**
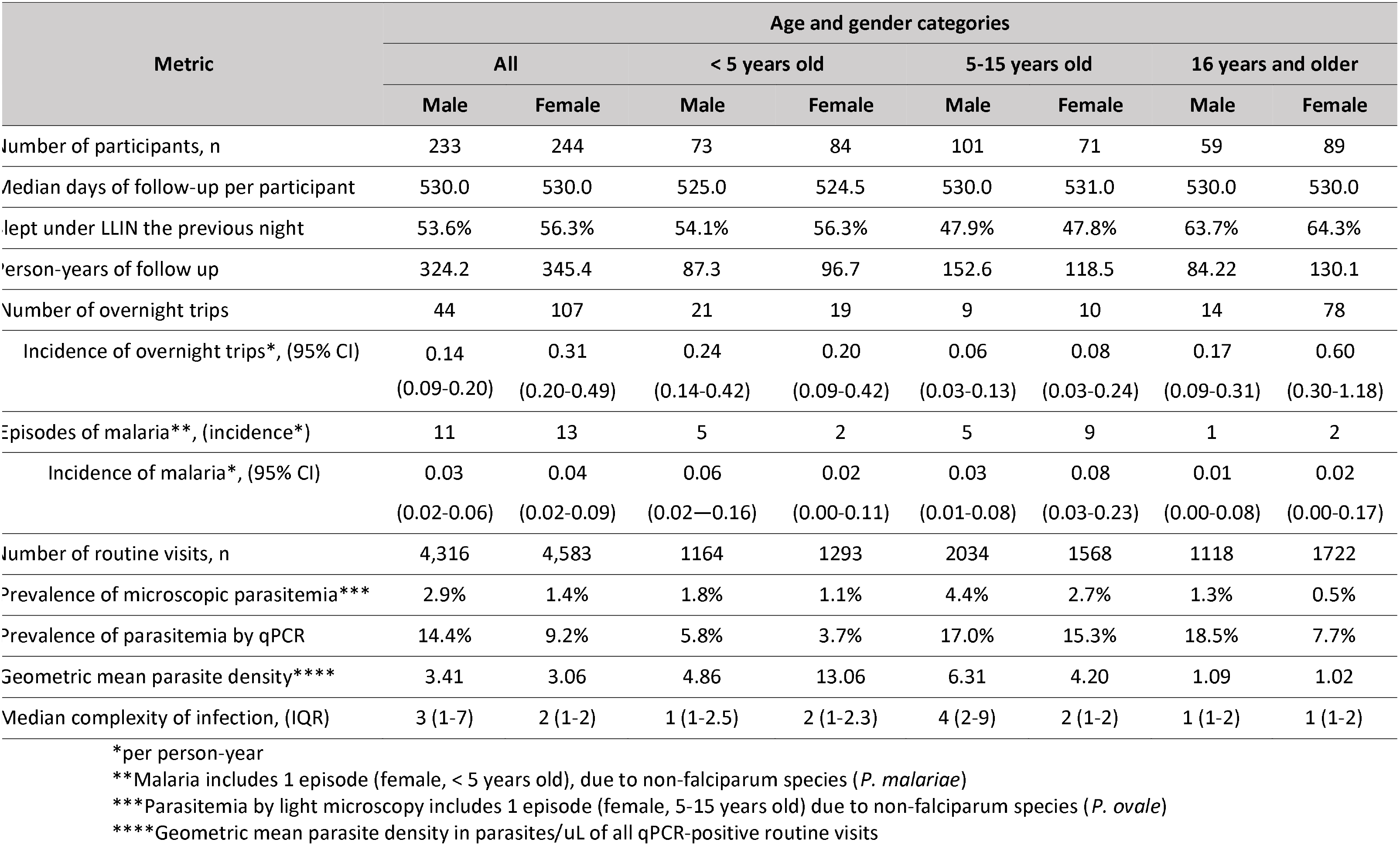
Behavioral risk factors for malaria infection and measures of malaria burden in study population, stratified by age and sex.

**Figure 1.**
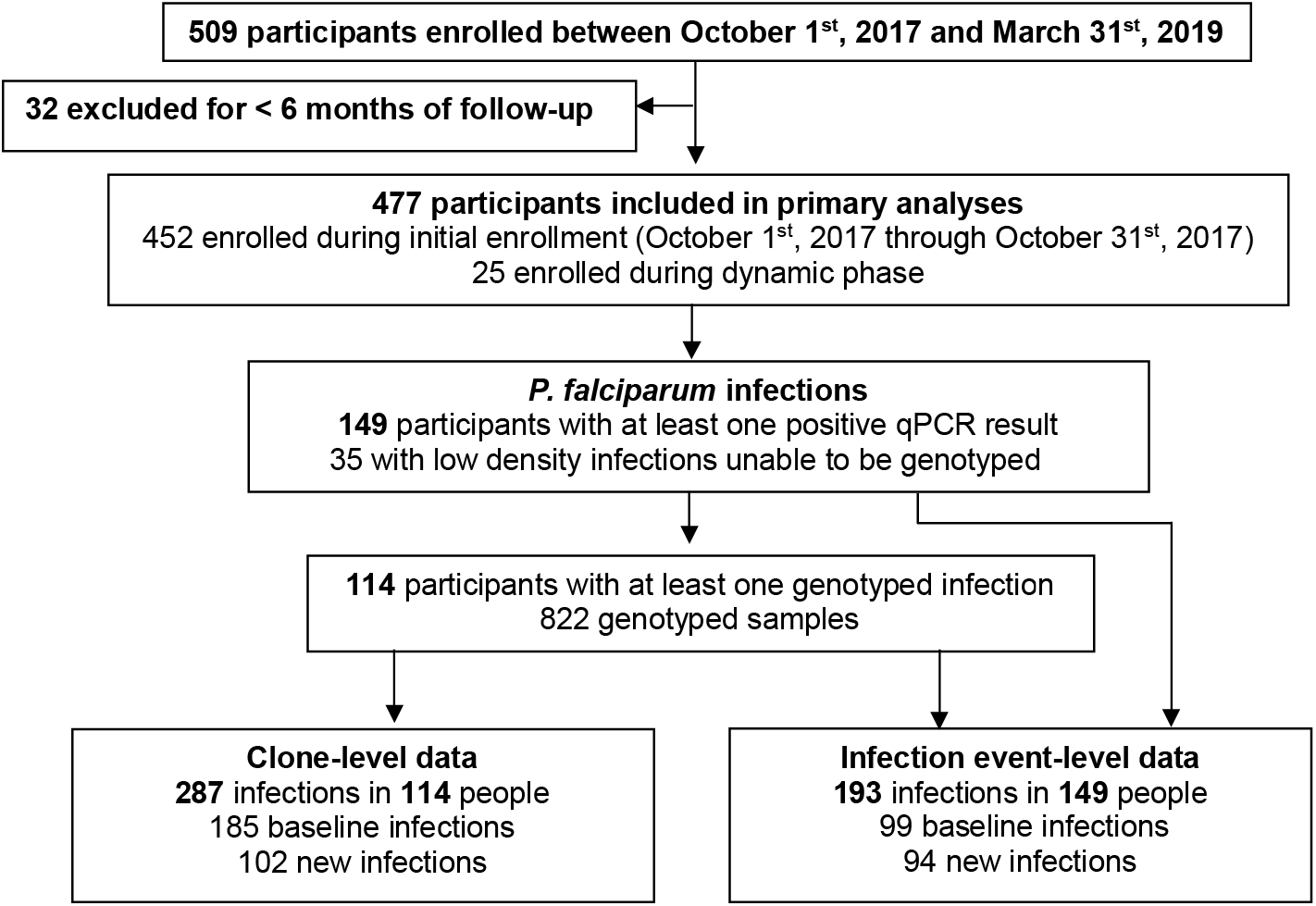
Study design.

### Behavioral malaria risk factors and measures of malaria burden

There was no difference in reported rates of LLIN use the previous night by sex (Table 1). Women over the age of 16 traveled overnight outside of the study area more than men (incidence rate ratio [IRR] for females vs. males = 3.61, 95% confidence interval [CI] 1.83 to 7.13), a potential risk factor for malaria exposure. Antimalarial use outside the study clinic was reported only 4 times (3 females and 1 male, all under the age of 5). In this region receiving regular rounds of IRS, the incidence of symptomatic malaria was low in all age categories, and there was no evidence of a difference in incidence of symptomatic malaria by sex overall (IRR for females vs. males = 1.11, 95% CI 0.49 to 2.52) or when adjusted for age (IRR = 1.26, 95% CI 0.54 to 2.96). In contrast, prevalence ratio (PR) of *P. falciparum* parasitemia by microscopy in females versus males across all age categories was 0.49 (95% CI 0.36 to 0.65), with relative differences in prevalence most pronounced in the oldest age group. Similar findings were seen when prevalence was assessed by ultrasensitive qPCR, with PR = 0.64 in females vs. males (95% CI 0.43 to 0.96), again with the largest differences seen in the oldest age group. Adjusting for age as a categorical variable, LLIN use, and travel did not qualitatively change prevalence ratios for microscopic parasitemia (PR in females vs. males = 0.57, 95% CI 0.42 to 0.77) or for qPCR-positive parasitemia (PR in females vs. males = 0.67, 95% CI 0.60 to 0.76). We also found no evidence for a difference in parasite density as determined by qPCR between males and females after adjusting for age as a continuous variable (p = 0.47). Median COI was higher in males than in females overall, driven primarily by a higher COI in male school aged children.

### Force of infection by age and sex

To determine whether higher infection prevalence in males was due to an increased rate of infection, we used longitudinal genotyping to calculate the force of infection (FOI, number of new blood stage infections per unit time). Overall, the FOI was low, with new infections occurring on average less than once every 5 years (Table 2). There was no evidence for a significant difference in FOI by sex overall (IRR for females vs. males = 0.88, 95% CI 0.48 to 1.62 by clone and IRR = 0.83, 95% CI 0.52 to 1.33 by infection event). There was also no evidence for a significant difference in FOI by sex when adjusted for age category (IRR = 0.88, 95% CI 0.47 to 1.63 by clone and IRR = 0.83, 95% CI 0.53 to 1.31 by infection event). For analysis both by clone and by infection event, there was a trend toward higher FOI in males compared to females. We performed a sensitivity analysis by decreasing the number of skips necessary to declare an infection cleared (Supplemental File 1f). Performing the same analysis using 1 skip for clearance instead of 3 skips increased the FOI in all groups and increased the trend toward higher FOI in males, with IRR for females vs. males when adjusted for age category = 0.71, 95% CI 0.41 to 1.24 by clone and IRR = 0.75, 95% CI 0.47 to 1.22 by infection event.

**Table 2.**
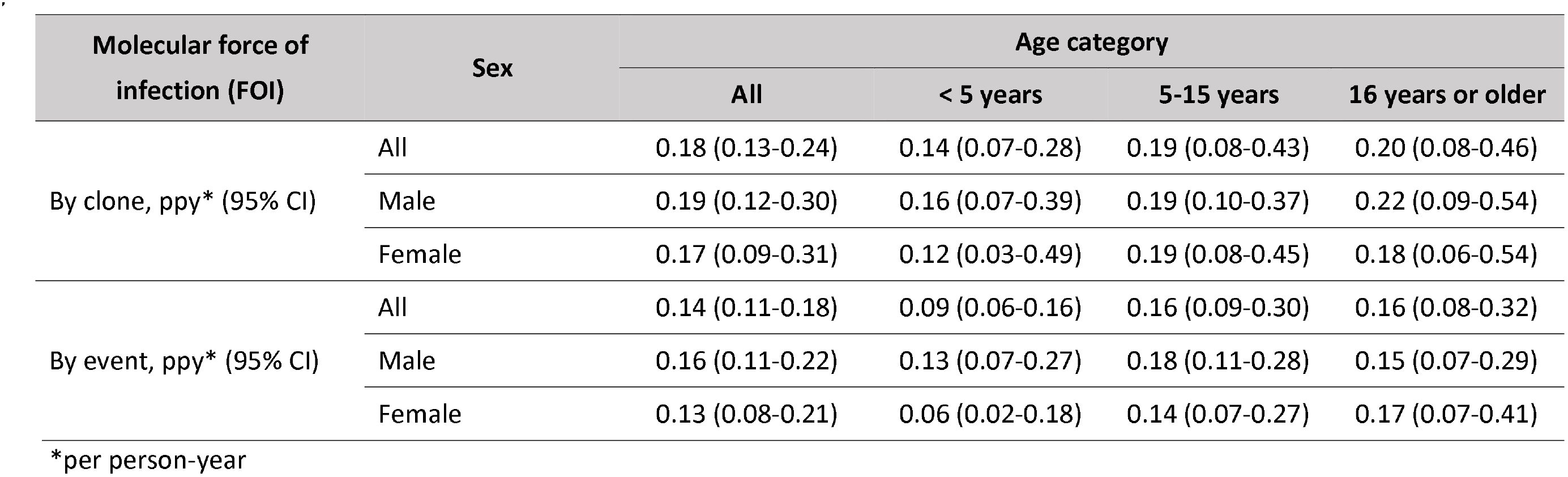
Molecular force of infection (FOI) by clone and by infection event, stratified by age and sex.

| Molecular force of<br>infection (FOI) | Sex | Age category |  |  |  |
| --- | --- | --- | --- | --- | --- |
|  |  | All | < 5 years | 5-15 years | 16 years or older |
| By clone, ppy* (95% CI) | All | 0.18 (0.13-0.24) | 0.14 (0.07-0.28) | 0.19 (0.08-0.43) | 0.20 (0.08-0.46) |
|  | Male | 0.19 (0.12-0.30) | 0.16 (0.07-0.39) | 0.19 (0.10-0.37) | 0.22 (0.09-0.54) |
|  | Female | 0.17 (0.09-0.31) | 0.12 (0.03-0.49) | 0.19 (0.08-0.45) | 0.18 (0.06-0.54) |
| By event, ppy* (95% CI) | All | 0.14 (0.11-0.18) | 0.09 (0.06-0.16) | 0.16 (0.09-0.30) | 0.16 (0.08-0.32) |
|  | Male | 0.16 (0.11-0.22) | 0.13 (0.07-0.27) | 0.18 (0.11-0.28) | 0.15 (0.07-0.29) |
|  | Female | 0.13 (0.08-0.21) | 0.06 (0.02-0.18) | 0.14 (0.07-0.27) | 0.17 (0.07-0.41) |

\*per person-year

### Rate of clearance of infection and duration of infection by sex

Since females had a lower prevalence of infection but similar rate of acquiring infections compared to males, we evaluated whether there was a difference between sexes in the rate at which infections were cleared. Asymptomatic infections were included in this analysis if they were not censored as stated in the methods; at the clone level, 105 baseline infections and 53 new infections were included. At the infection event level, 58 baseline infections and 51 new infections were included. There were more baseline infections in males at the clone level, with 68/105 (64.8%) baseline infections in males and 37/105 (35.2%) baseline infections in females (p <0.001). At the infection event level, there was no difference in the number of baseline infections by sex, with 32/58 (55.2%) baseline infections in males and 26/58 (44.8%) baseline infections in females (p = 0.35).

Unadjusted hazard ratios for clearing infecting clones showed that asymptomatic infections cleared naturally *(i.e*., when not treated by antimalarials) at nearly twice the rate in females vs. males (hazard ratio (HR) 1.92, 95% CI 1.19 to 3.11, Table 3). In addition, new infections cleared faster than baseline infections and monoclonal infections cleared faster than polyclonal infections. Unadjusted hazard ratios for clearance of infection events (as opposed to clones) also showed faster clearance in females vs. males (HR = 2.30, 95% CI 1.20 to 4.42). Results were similar in multivariate models including age, gender, the period during which the infection was first observed, and parasite density, demonstrating faster clearance in females vs. males (HR =1.82, 95% CI 1.20 to 2.75 by clone and HR =2.07, 95% CI 1.24 to 3.47 by infection event). Complexity of infection was not included in the final adjusted model because the model fit the data less well when COI was included as a predictor. In both adjusted models, new infections cleared faster than baseline infections. Higher parasite densities were associated with slower clearance by clone and by infection event, but the effect size was larger when data were analyzed by infection event (HR = 0.44, 95% CI 0.35 to 0.54). There was no evidence for interaction between age and sex in either adjusted model. We performed a sensitivity analysis by decreasing the number of skips necessary to declare an infection cleared. Regardless of whether 3 skips, 2 skips, or 1 skip was used to determine infection clearance, females cleared their infections faster than males both by clone and by infection event (Supplemental File 1g).

**Table 3.** Hazard ratios for rates of clearance of infection, by clone and by infection event.

| Predictors | Categories | Hazard ratio by clone (95% CI) |  | Hazard ratio by infection event (95% CI) |  |
| --- | --- | --- | --- | --- | --- |
|  |  | Unadjusted | Adjusted | Unadjusted | Adjusted |
| Sex | Male | ref | ref | ref | ref |
|  | Female | 1.92 (1.19-3.11) | 1.82 (1.20 – 2.75) | 2.30 (1.20 – 4.42) | 2.07 (1.24 – 3.47) |
| Age | 16 years or greater | ref | ref | ref | ref |
|  | 5-15 years | 0.66 (0.39 – 1.10) | 0.81 (0.49 – 1.36) | 0.82 (0.39 – 1.74) | 1.27 (0.72 – 2.25) |
|  | < 5 years | 1.64 (0.79 – 3.41) | 1.55 (0.76 – 3.17) | 2.01 (0.80 – 5.00) | 1.75 (0.87 – 3.53) |
| Complexity of infection (COI) | Polyclonal (COI > 1) | ref | -- | ref | -- |
|  | Monoclonal (COI = 1) | 1.63 (1.03 – 2.57) | -- | 0.95 (0.38 – 2.34) | -- |
| Infection status | Present at baseline | ref | ref | ref | ref |
|  | New infection | 1.94 (1.22 – 3.07) | 1.75 (1.05 – 2.94) | 4.66 (2.58 – 8.42) | 4.32 (2.59 – 7.20) |
| Parasite density * |  | 0.85 (0.69 – 1.06) | 0.81 (0.65 – 1.00) | 0.41 (0.32 – 0.51) | 0.44 (0.35 – 0.54) |

\*Increasing parasite density (log10) in parasites/microliter, as measured by qPCR

We next estimated durations of asymptomatic infection by age and sex using results from a model that included these covariates (Figure 2). Durations of infection ranged from 103 days to 447 days by clone, and from 87 to 536 days by infection event. Males had a longer duration of infection across all age categories. Children aged 5-15 years had the longest duration of infection, followed by adults. Therefore, overall, males aged 5-15 years had the longest estimated duration of infection by either clone (447 days) or infection event (526 days).

**Figure 2.**
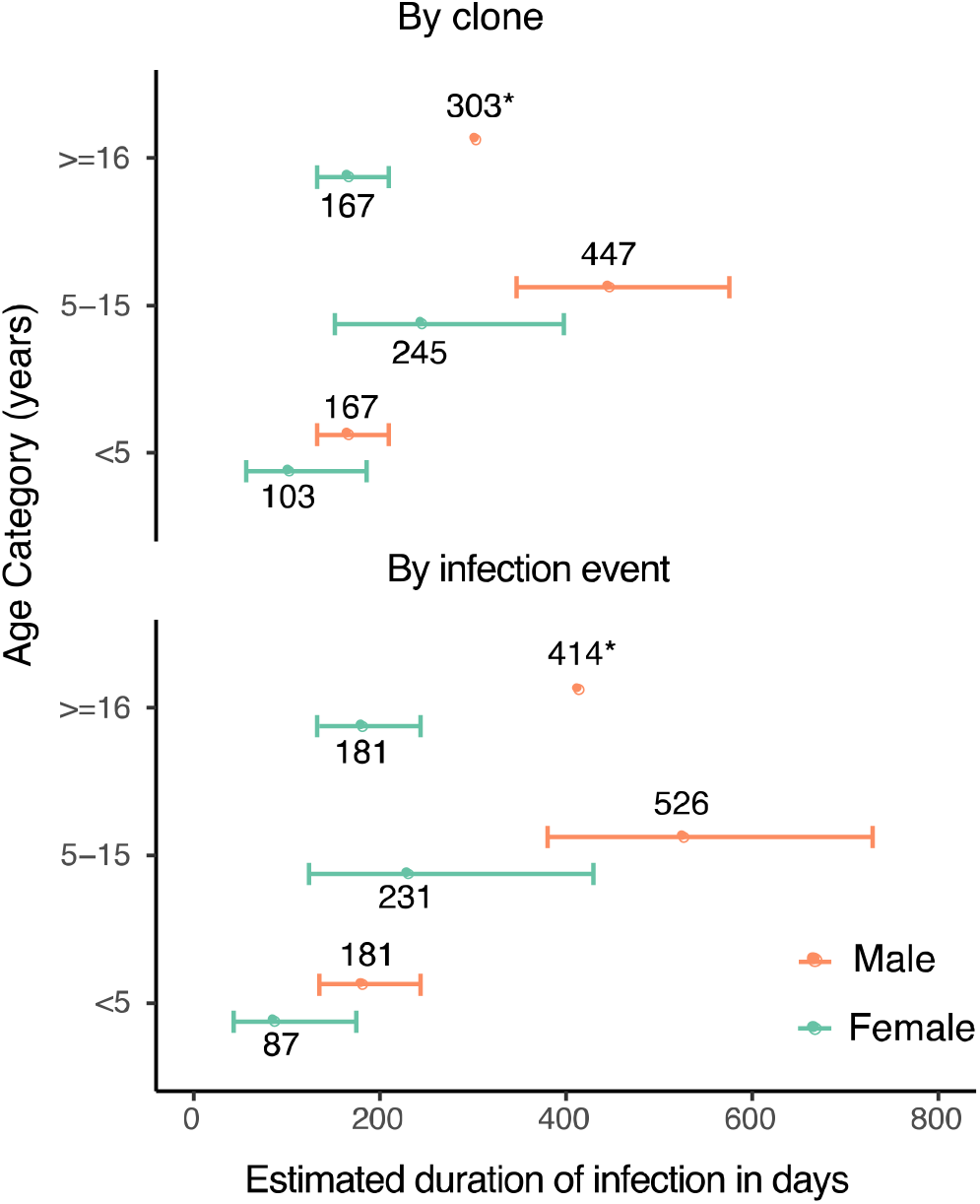
Estimates of duration of infection from sex- and age-adjusted model. Estimated duration of infection in days, calculated by adjusting the point estimate of the baseline hazard by the coefficients of the sex- and age-adjusted model. Error bars represent standard errors of duration obtained from variance in the model coefficients. Point estimates of duration are labeled (*).

## Discussion

Previous studies have reported a higher prevalence of malaria infection in males compared to females, with the difference often ascribed to differences in exposure^10-16^. In a cohort study in eastern Uganda, we noted higher prevalence of malaria infection in males compared to females. By closely following a cohort of children and adults and genotyping every detected infection with sensitive amplicon deep-sequencing, we were able to estimate both the rate of infection (FOI) and duration of infection and to compare these measures by sex. We found that lower prevalence in females did not appear to be due to lower rates of infection but rather due to faster clearance of asymptomatic infections. To our knowledge, this is the first study to report a sex-based difference in the duration of malaria infection.

Many prior studies of malaria incidence and/or prevalence that evaluated associations with sex in late childhood, adolescence and adulthood have found a male bias in the observed measure of burden^10-16^. Overall, these studies consistently suggest that males exhibit higher incidence and/or prevalence of malaria that begins during late childhood, persisting through puberty and the majority of adulthood (excepting the years when pregnancy puts women at higher risk). One possible explanation put forward for the sex-specific difference in burden has been that males are more frequently bitten by malaria-carrying mosquitos due to behavioral differences such as working outside, not sleeping under a net, or traveling for work^11,13,17,18^. In our study, however, there were no statistically significant differences in malaria incidence or FOI by sex, though there was a trend toward higher FOI in males. We also saw no evidence of behavioral trends that would result in more infections in males; in fact, older women in our study did most of the traveling outside the study area (an area of low transmission compared to surrounding areas). We did not assess work habits as part of our study questionnaire, but the fact that we observe similar patterns in prevalence by sex in all age categories makes this a less likely explanation. Therefore, in this particular cohort, it appears that the observed male bias in parasite prevalence is best explained by slower clearance of infection in males. We cannot rule out that males were differentially exposed in the recent past (e.g., had a higher force of infection when transmission was higher, which could have affected immunity), and we plan to genotype samples from a prior cohort from 2011-2017 to test this hypothesis.

Very few studies have been conducted to explore immunological differences between males and females in their response to the malaria parasite. RTS,S vaccination is associated with higher all-cause mortality in girls compared to boys, and a trend toward higher risk of fatal malaria has been noted in vaccinated girls compared to boys, suggesting possible sexual dimorphism in immunological responses to malaria^23^. The comprehensive Garki project found that females had higher levels of antibodies against *P. falciparum* compared to men^10^. Hormonal differences have been posited to play a role in these sex-based immunological differences to malaria infection; studies in mice show that testosterone appears to downregulate the immune response to malaria^24,25^. If sex-based differences were purely hormonally mediated, one might expect a stronger effect in adults than in school-aged children, which was not seen in this cohort. This could be due to imperfect detectability, as adults have lower density infections than school-aged children due to improved anti-parasite immunity^2,3^. Post-menopausal women were also included in the oldest age group, which could have blunted the effect of sex in that age group if there is a hormonal basis for some of this effect. It is also possible that sex-based differences are explained by a different mechanism, such as differences in innate immunity. More studies are needed to elucidate the relationship between sex-based biological differences between males and females and their impact on the development of effective antimalarial immunity in humans.

Of the variables we evaluated in addition to sex, baseline infection status and parasite density were most strongly associated with the rate of clearance of infection. Infections that were already present at the beginning of the study and persisted past the left censoring date may have been a non-random selection of well-established asymptomatic infections that were present at baseline at a higher frequency than average because they had a fundamentally different trajectory than newly established asymptomatic infections. Higher parasite densities were also associated with slower clearance of infection in both adjusted models, but the effect was most pronounced when the data were analyzed by infection event. This may be because low-density infections that we were unable to successfully genotype were only included in the infection event analysis, and these events tended to have short durations. The inclusion of parasite density in our multivariate models did not meaningfully alter associations between sex and duration of infection, providing evidence that the sex-based differences in duration were not mediated primarily by differences in parasite density in our cohort.

A limitation of our study is the statistical model’s assumption that all infections clear at the same rate, which was necessary because we did not observe the beginning of most infections. To rely less on this assumption, we adjusted for baseline infections, allowing them to have a different rate of clearance than new infections; this did not change our primary finding of a difference in rate of clearance by sex. We also acknowledge there may be other unmeasured confounders, such as genetic hemoglobinopathies or sex-based differences in unreported outside antimalarial use, that could affect our results. Another caveat is that our findings may not be generalizable to areas with different transmission intensity given that our study was conducted in a unique area with previously very high transmission intensity that has been greatly reduced in recent years by repeated rounds of IRS.

Additionally, because it is difficult to genotype low density infections and because parasite densities fluctuate over time, we allowed several "skips” in detection before declaring an infection cleared or the same clone in an individual a new infection with the same clone. This requires the assumption that re-infection with the same clone is relatively rare, which is reasonable given the high genetic diversity and low force of infection seen in this setting; this assumption has been made in other longitudinal genotyping studies^26,27^. Our method may have biased us toward longer durations of infection and fewer new infections overall, but this is unlikely to have introduced any significant bias in the observed associations by sex. We performed a sensitivity analysis to ensure that changing the number of skips allowed did not significantly change the main finding regarding longer duration of infection in males and our findings were consistent. Given decreasing parasite densities over time and a setting with low force of infection and low EIR, assuming perfect recovery of clones is likely to artificially inflate the force of infection in this cohort.

In summary, we estimated the clearance of asymptomatic *P. falciparum* infections by genotyping longitudinal samples from a cohort in Nagongera, Uganda, using sensitive amplicon deep-sequencing and found that females cleared their infections at a faster rate than males; this finding remained consistent when adjusting for age, baseline infection status, and parasite density. Furthermore, we found no conclusive evidence for a sex-based difference in exposure to infection, either behaviorally or by FOI. Though there have long been observed differences in malaria burden between the sexes, there is still little known about sex-based biological differences that may mediate immunity to malaria. Unfortunately, much reported malaria data is still sex-disaggregated. Our findings should encourage epidemiologists to better characterize sexual dimorphism in malaria and motivate increased research into biological explanations for sex-based differences in the host response to the malaria parasite.

## Materials and Methods

### Study setting and population-level malaria control interventions

This cohort study was carried out in Nagongera sub-county, Tororo district, eastern Uganda, an area with historically high malaria transmission. However, 7 rounds of indoor residual spraying (IRS) from 2014-2019 have resulted in a significant decline in the burden of malaria^28^. Pre-IRS, the daily human biting rate (HBR) was 34.3 and the annual entomological inoculation rate (EIR) was 238; after 5 years of IRS, in 2019 the daily HBR was 2.07 and overall annual EIR was 0.43 as reported by Nankabirwa et. al.^29^

### Study design, enrollment, and follow-up

All members of 80 randomly selected households with at least two children were enrolled in October 2017 using a list generated by enumerating and mapping all households in Nagongera sub-county^29^. The cohort was dynamic such that residents joining the household were enrolled and residents leaving the household were withdrawn. Data for this analysis was collected from October 1^st^, 2017 through March 31^st^, 2019; participants were included if they had at least 6 months of contiguous follow up. Participants were followed at a designated study clinic open daily from 8 AM to 5 PM. Participants were encouraged to seek all medical care at the study clinic and avoid the use of antimalarial medications outside of the study. Routine visits were conducted every 28 days and included a standardized clinical evaluation, assessment of overnight travel outside of Nagongera sub-county, and collection of blood by phlebotomy for detection of malaria parasites by microscopy and molecular studies. Participants came in for non-routine visits in the setting of illness. Blood smears were performed at enrollment, at all routine visits and at non-routine visits if the participant presented with fever or history of fever in the previous 24 hours. Participants with fever (>38.0°C tympanic) or history of fever in the previous 24 hours had a thick blood smear read urgently. If the smear was positive, the patient was diagnosed with malaria and treated with artemether-lumefantrine. Participants with asymptomatic parasitemia as detected by qPCR or microscopy were not treated with antimalarials, consistent with Uganda national guidelines. Study participants were visited at home every 2 weeks to assess use of long-lasting insecticidal nets (LLINs) the previous night.

### Laboratory methods

Thick blood smears were stained with 2% Giemsa for 30 minutes and evaluated for the presence of asexual parasites and gametocytes. Parasite densities were calculated by counting the number of asexual parasites per 200 leukocytes (or per 500, if the count was less than 10 parasites per 200 leukocytes), assuming a leukocyte count of 8,000/μl. A thick blood smear was considered negative if examination of 100 high power fields revealed no asexual parasites. For quality control, all slides were read by a second microscopist, and a third reviewer settled any discrepant readings. In our experienced microscopists’ hands, the lower limit of detection is approximately 20-50 parasites/µL.

For qPCR and genotyping, we collected 200 µL of blood at enrollment, at each routine visit, and at the time of malaria diagnosis. DNA was extracted using the PureLink Genomic DNA Mini Kit (Invitrogen) and parasitemia was quantified using an ultrasensitive varATS qPCR assay with a lower limit of detection of 0.05 parasites/µL^30^. Samples with a parasite density >= 0.1 parasites/µL blood were genotyped via amplicon deep-sequencing. All samples positive for asexual parasites by microscopy but negative for *P. falciparum* by qPCR were tested for the presence of non-falciparum species using nested PCR.^31^

#### Sequencing library preparation

Hemi-nested PCR was used to amplify a 236 base-pair segment of apical membrane antigen 1 (AMA-1) using a published protocol^32^, with modifications **(Supplemental File 1a)**. Samples were amplified in duplicate, indexed, pooled, and purified by bead cleaning. Sequencing was performed on an Illumina MiSeq platform (250bp paired-end).

### Bioinformatics methods

Data extraction, processing, and haplotype clustering were performed using SeekDeep^33^, followed by additional filtering^34^. **Supplemental File 1b** shows the full bioinformatics workflow.

## Data Availability

All data is available by request from the authors. After acceptance and publication by a peer-reviewed journal, all data will be published on ClinEpiDB.org.

## Data analysis

A clone was defined as a genetically identical group of parasites, e.g., with identical haplotypes. Because polyclonal infections can occur due to co-infection (one mosquito bite transmitting multiple clones) or superinfection (multiple bites), we analyzed the infection data both by clone and by infection event. For analysis by clone, each unique clone was counted as an infection and each clone’s disappearance as a clearance event. For analysis by infection event, any new clones seen within 3 visits of the date of the first newly detected clone(s) were grouped together and considered one new "infection event.” Clearance of infection for these events required that all clones in the group be absent. A baseline infection was defined as a clone or infection event (group of clones) detected in the first 60 days of observation. New infections were defined as a new clone or infection event (group of clones) detected in an individual after day 60.

Imperfect detection of *P. falciparum* clones is known to be a limitation of all PCR-based genotyping methods; both biological factors, such as deep organ sequestration of clones in the lifecycle of the parasite, and methodological factors, such as difficulty amplifying minority clones (especially at lower parasite densities), results in imperfect detectability of all clones present in a single blood sample drawn on a single day.^26,35-38^ Though amplicon deep sequencing is less likely than other methods to miss minority clones^39^, detectability was a particular concern in our study, in which the majority of infections were submicroscopic and parasite densities declined over time [Supplemental File 1c]. To account for the fact that a clone might be missed in a single sample due to fluctuations in parasite density and/or methodological limitations, we allowed 3 "skips” in detection and classified an infection as cleared only if it was not identified in 4 contiguous samples from routine visits. If the infection (as determined by clone or by infection event) was absent for 4 routine samples in a row, the last date of detection would be the end date of that infection. Therefore, for a clone that had previously infected an individual to be classified as a new infection, it had to have been absent from that individual for at least 4 routine visits. Additional details are found in **Supplemental File 1d**. This decision was supported by the fact that the diversity of the genotyped AMA-1 amplicon was quite high (expected heterozygosity = 0.949), resulting in a low probability (5.1%) for infection with the same clone by chance.

Data analysis was conducted in R^40^ and Python^41^. Microscopic parasite prevalence was defined by the number of smear-positive routine visits over all routine visits. Parasite prevalence by qPCR was defined as the number of qPCR-positive visits over all routine visits. Prevalence ratios were computed using Poisson regression with generalized estimating equations to adjust for repeated measures. Comparison of parasite density by sex was made using linear regression with generalized estimating equations to adjust for repeated measures. Force of infection (FOI) was defined as the number of new infections, including malaria episodes, divided by person time. Poisson regression with generalized estimating equations to account for repeated measures was used to estimate malaria incidence, FOI, and to calculate incident rate ratios (IRR) for malaria and FOI. Hazards for clearance of untreated, asymptomatic infections were estimated using time-to-event models (shared frailty models fit using R package “frailtypack,” version 2.12.2)^42,43^. Infections were censored if they were only observed in the first three months or the last three months (before January 01, 2018 and after January 01, 2019) because they were not observed for long enough to determine whether clearance occurred. If an infection was observed for only one timepoint, it was assigned a duration of 14 days. These models assumed a constant hazard of clearance and included random effects to account for repeated measures in individuals. Parasite density was included in the model as a time-varying covariate. Duration of infection in days was calculated as 1/adjusted hazard.

## Data Accessibility

Data from the cohort study is available through an open-access clinical epidemiology database resource, ClinEpiDB at https://clinepidb.org/ce/app/record/dataset/DS_51b40fe2e2. Genotyping data and code used to generate tables and figures is available on GitHub^44^.

## Supplemental File Legends

**Supplemental File 1a. AMA-1 hemi-nested PCR protocol for amplicon deep-sequencing**

**Supplemental File 1b: Bioinformatics workflow**

**Supplemental File 1c. Declining qPCR density over time in the cohort**.

**Supplemental File 1d. Detailed explanation of skip rule criteria**

**Supplemental File 1e. Haplotype sequences and frequencies**

**Supplemental File 1f. Sensitivity analysis of molecular force of infection: Table 2 replicated using 2 skips or 1 skip**

**Supplemental File 1g. Sensitivity analysis of duration of infection: Table 3 replicated using 2 skips or 1 skip**

## Notes

### Competing Interest Statement

The authors have declared no competing interest.

### Funding Statement

Funding was provided by the National Institutes of Health as part of the International Centers of Excellence in Malaria Research (ICMER) program (U19AI089674). JB is supported by the National Institutes of Health training grant T32 AI007641-16. JIN is supported by the Fogarty International Center (Emerging Global Leader Award grant number K43TW010365). EA is supported by the Fogarty International Center of the National Institutes of Health under Award Number D43TW010526. The ClinEpiDB platform is supported by award OPP1169785 from the Bill & Melinda Gates Foundation. The funders had no role in the study
design, data collection and analysis, decision to publish or preparation of the manuscript.

### Author Declarations

This study protocol was approved by both the UCSF IRB and the UNCST IRB in Uganda.

## REFERENCES

1. World malaria report 2019 [Internet], [cited 2020 Aug 5]. Available from: https://www.who.int/publications-detail-redirect/9789241565721

2. Bousema T, Okell L, Felger I, Drakeley C. Asymptomatic malaria infections: detectability, transmissibility and public health relevance. Nat Rev Microbiol. 2014;12(12):833–840. PMID: 25329408

3. Okell LC, Bousema T, Griffin JT, Ouédraogo AL, Ghani AC, Drakeley CJ. Factors determining the occurrence of submicroscopic malaria infections and their relevance for control. Nature Communications. 2012 Dec 4;3(1):1-9.

4. Tadesse FG, Slater HC, Chali W, Teelen K, Lanke K, Belachew M, Menberu T, Shumie G, Shitaye G, Okell LC, Graumans W, van Gemert G-J, Kedir S, Tesfaye A, Belachew F, Abebe W, Mamo H, Sauerwein R, Balcha T, Aseffa A, Yewhalaw D, Gadisa E, Drakeley C, Bousema T. The Relative Contribution of Symptomatic and Asymptomatic Plasmodium vivax and Plasmodium falciparum Infections to the Infectious Reservoir in a Low-Endemic Setting in Ethiopia. Clin Infect Dis. 2018 01;66(12):1883–1891. PMID: 29304258

5. Slater HC, Ross A, Felger I, Hofmann NE, Robinson L, Cook J, Gonçalves BP, Björkman A, Ouedraogo AL, Morris U, Msellem M, Koepfli C, Mueller I, Tadesse F, Gadisa E, Das S, Domingo G, Kapulu M, Midega J, Owusu-Agyei S, Nabet C, Piarroux R, Doumbo O, Doumbo SN, Koram K, Lucchi N, Udhayakumar V, Mosha J, Tiono A, Chandramohan D, Gosling R, Mwingira F, Sauerwein R, Riley EM, White NJ, Nosten F, Imwong M, Bousema T, Drakeley C, Okell LC. The temporal dynamics and infectiousness of subpatent Plasmodium falciparum infections in relation to parasite density. Nature Communications. 2019 Mar 29;10(1):1433.

6. Zuk M, McKean KA. Sex differences in parasite infections: Patterns and processes. International Journal for Parasitology. 1996 Oct 1;26(10):1009–1024.

7. Klein SL. Hormonal and immunological mechanisms mediating sex differences in parasite infection. Parasite Immunology. 2004;26(6-7):247-264.

8. Roberts CW, Walker W, Alexander J. Sex-Associated Hormones and Immunity to Protozoan Parasites. Clin Microbiol Rev. 2001 Jul 1; 14(3):476-488.

9. Desai M, Kuile FO ter, Nosten F, McGready R, Asamoa K, Brabin B, Newman RD. Epidemiology and burden of malaria in pregnancy. The Lancet Infectious Diseases. 2007 Feb 1;7(2):93–104. PMID: 17251080

10. Molineaux L, Gramiccia G, Organization WH. The Garki projectµ: research on the epidemiology and control of malaria in the Sudan savanna of West Africa [Internet]. WorldHealth Organization; 1980 [cited 2020 Feb 18]. Available from: https://apps.who.int/iris/handle/10665/40316

11. Pathak S, Rege M, Gogtay NJ, Aigal U, Sharma SK, Valecha N, Bhanot G, Kshirsagar IMA, Sharma S. Age-Dependent Sex Bias in Clinical Malarial Disease in Hypoendemic Regions. PLoS One [Internet]. 2012 Apr 25 [cited 2020 Feb 18];7(4). Available from: https://www.ncbi.nlm.nih.gov/pmc/articles/PMC3338423/ PMCID: PMC3338423

12. Landgraf B, Kollaritsch H, Wiedermann G, Wernsdorfer WH. Parasite density of Plasmodium falciparum malaria in Ghanaian schoolchildren: evidence for influence of sex hormones? Transactions of the Royal Society of Tropical Medicine and Hygiene. 1994 Jan;88(1):73-74.

13. Camargo LMA, Colletto GMDD, Ferreira MU, Gurgel SDM, Escobar AL, Marques A, Krieger H, Camargo EP, Silva LHPD. Hypoendemic Malaria in Rondonia (Brazil, Western Amazon Region): Seasonal Variation and Risk Groups in an Urban Locality. The American Journal of Tropical Medicine and Hygiene. 1996 Jul 1;55(1):32-38.

14. Abdalla SI, Malik EM, Ali KM. The burden of malaria in Sudan: incidence, mortality and disability-adjusted life-years. Malar J. 2007 Jul 28;6:97. PMCID: PMC1995207

15. Houngbedji CA, N’Dri PB, Hürlimann E, Yapi RB, Silué KD, Soro G, Koudou BG, Acka CA, Assi S-B, Vounatsou P, N’Goran EK, Fantodji A, Utzinger J, Raso G. Disparities of Plasmodium falciparum infection, malaria-related morbidity and access to malaria prevention and treatment among school-aged children: a national cross-sectional survey in Côte d’Ivoire. Malar J [Internet]. 2015 Jan 5 [cited 2020 Aug 5];14. Available from: https://www.ncbi.nlm.nih.gov/pmc/articles/PMC4326184/ PMCID: PMC4326184

16. Mulu A, Legesse M, Erko B, Belyhun Y, Nugussie D, Shimelis T, Kassu A, Elias D, Moges B. Epidemiological and clinical correlates of malaria-helminth co-infections in southern Ethiopia. Malar J. 2013 Jul 3;12:227. PMCID: PMC3706225

17. Moon JJ, Cho S-Y. Incidence patterns of vivax malaria in civilians residing in a high-risk county of Kyonggi-do (Province), Republic of Korea. Korean J Parasitol. 2001 Dec;39(4):293-299. PMCID: PMC2721214

18. Finda MF, Moshi IR, Monroe A, Limwagu AJ, Nyoni AP, Swai JK, Ngowo HS, Minja EG, Toe LP, Kaindoa EW, Coetzee M, Manderson L, Okumu FO. Linking human behaviours and malaria vector biting risk in south-eastern Tanzania. PLoS One [Internet]. 2019 Jun 3 [cited 2020 Apr 8];14(6). Available from: https://www.ncbi.nlm.nih.gov/pmc/articles/PMC6546273/ PMCID: PMC6546273

19. Nhamoyebonde S, Leslie A. Biological Differences Between the Sexes and Susceptibility to Tuberculosis. J Infect Dis. 2014 Jul 15;209(suppl_3):S100-S106.

20. Bernin H, Lotter H. Sex Bias in the Outcome of Human Tropical Infectious Diseases: Influence of Steroid Hormones. J Infect Dis. 2014 Jul 15;209(suppl_3):S107-S113.

21. Fischer J, Jung N, Robinson N, Lehmann C. Sex differences in immune responses to infectious diseases. Infection. 2015 Aug 1;43(4):399-403.

22. Fish EN. The X-files in immunity: sex-based differences predispose immune responses. Nat Rev Immunol. 2008 Sep;8(9):737-744.

23. Klein SL, Shann F, Moss WJ, Benn CS, Aaby P. RTS,S Malaria Vaccine and Increased Mortality in Girls. mBio [Internet]. 2016 Apr 26 [cited 2020 Feb 18];7(2). Available from: https://www.ncbi.nlm.nih.gov/pmc/articles/PMC4850267/ PMCID: PMC4850267

24. Delic D, Ellinger-Ziegelbauer H, Vohr HW, Dkhil M, Al-Quraishy S, Wunderlich F. Testosterone response of hepatic gene expression in female mice having acquired testosterone-unresponsive immunity to Plasmodium chabaudi malaria. Steroids. 2011 Oct;76(10-11): 1204-1212. PMID: 21669218

25. Wunderlich, F, Marinovski P, Peter W, Benten M, Schmitt-Wrede, H-P, Mossman H. Testosterone and Other Gonadal Factor(s) Restrict the Efficacy of Genes Controlling Resistance to Plasmodium Chabaudi Malaria [Internet]. Parasite immunology. 1991. Parasite Immunology, 13: 357-367. doi:10.1111/j.1365-3024.1991.tb00289.x

26. Feiger I, Maire M, Bretscher MT, Falk N, Tiaden A, Sama W, Beck H-P, Owusu-Agyei S, Smith TA. The Dynamics of Natural Plasmodium falciparum Infections. PLOS ONE. 2012 Sep 18;7(9):e45542.

27. Smith T, Felger I, Fraser-Hurt N, Beck H-P. 10. Effect of insecticide-treated bed nets on the dynamics of multiple Plasmodium falciparum infections. Transactions of the Royal Society of Tropical Medicine and Hygiene. 1999 Feb 1;93:53-57.

28. Nankabirwa JI, Briggs J, Rek J, Arinaitwe E, Nayebare P, Katrak S, Staedke SG, Rosenthal PJ, Rodriguez-Barraquer I, Kamya MR, Dorsey G, Greenhouse B. Persistent Parasitemia Despite Dramatic Reduction in Malaria Incidence After 3 Rounds of Indoor Residual Spraying in Tororo, Uganda. J Infect Dis. 2019 Mar 15;219(7):1104–1111. PMCID: PMC6420168

29. Nankabirwa JI, Arinaitwe E, Rek J, Kilama M, Kizza T, Staedke SG, Rosenthal PJ, Rodriguez-Barraquer I, Briggs J, Greenhouse B, Bousema T, Drakeley C, Roos DS, Tomko SS, Smith DL, Kamya MR, Dorsey G. Malaria Transmission, Infection, and Disease following Sustained Indoor Residual Spraying of Insecticide in Tororo, Uganda. The American Journal of Tropical Medicine and Hygiene; 2020 Jul 20;tpmd200250.

30. Hofmann N, Mwingira F, Shekalaghe S, Robinson LJ, Mueller I, Felger I. Ultra-sensitive detection of Plasmodium falciparum by amplification of multi-copy subtelomeric targets. PLoS Med. 2015 Mar;12(3):el001788. PMCID: PMC4348198

31. Snounou G, Viriyakosol S, Zhu XP, Jarra W, Pinheiro L, do Rosario VE, Thaithong S, Brown KN. High sensitivity of detection of human malaria parasites by the use of nested polymerase chain reaction. Mol Biochem Parasitol. 1993 oct;61(2):315-320. PMID: 8264734

32. Miller RH, Hathaway NJ, Kharabora O, Mwandagalirwa K, Tshefu A, Meshnick SR, Taylor SM, Juliano JJ, Stewart VA, Bailey JA. A deep sequencing approach to estimate Plasmodium falciparum complexity of infection (COI) and explore apical membrane antigen 1 diversity. Malar J. 2017 Dec 16;16(1):490. PMCID: PMC5732508

33. Hathaway NJ, Parobek CM, Juliano JJ, Bailey JA. SeekDeep: single-base resolution de novo clustering for amplicon deep sequencing. Nucleic Acids Res. 2018 Feb 28;46(4):e21-e21.

34. Lerch A, Koepfli C, Hofmann NE, Kattenberg JH, Rosanas-Urgell A, Betuela I, Mueller I, Feiger I. Longitudinal tracking and quantification of individual Plasmodium falciparum clones in complex infections. Sci Rep [Internet]. 2019 Mar 4 [cited 2020 Feb 18];9. Available from: https://www.ncbi.nlm.nih.gov/pmc/articles/PMC6399284/ PMCID: PMC6399284

35. Koepfli C, Mueller I. Malaria Epidemiology at the Clone Level. Trends Parasitol. 2017 Dec;33(12):974-985. PMCID: PMC5777529

36. Nguyen T-N, von Seidlein L, Nguyen T-V, Truong P-N, Hung SD, Pham H-T, Nguyen T-U, Le TD, Dao VH, Mukaka M, Day NP, White NJ, Dondorp AM, Thwaites GE, Hien TT. The persistence and oscillations of submicroscopic Plasmodium falciparum and Plasmodium vivax infections overtime in Vietnam: an open cohort study. Lancet Infect Dis. 2018 May;18(5):565-572. PMCID: PMC5910058

37. Koepfli C, Schoepflin S, Bretscher M, Lin E, Kiniboro B, Zimmerman PA, Siba P, Smith TA, Mueller I, Felger I. How much remains undetected? Probability of molecular detection of human Plasmodia in the field. PLoS ONE. 2011 Apr 28;6(4):e19010. PMCID: PMC3084249

38. Ross A, Koepfli C, Li X, Schoepflin S, Siba P, Mueller I, Felger I, Smith T. Estimating the numbers of malaria infections in blood samples using high-resolution genotyping data. PLoS ONE. 2012;7(8):e42496. PMCID: PMC3430681

39. Lerch A, Koepfli C, Hofmann NE, Messerli C, Wilcox S, Kattenberg JH, Betuela I, O’Connor L, Mueller I, Felger I. Development of amplicon deep sequencing markers and data analysis pipeline for genotyping multi-clonal malaria infections. BMC Genomics. 2017 Nov 13;18(1):864.

40. R Core Team. R: A language and environment for statistical computing. [Internet]. R Foundation for Statistical Computing, Vienna, Austria.; 2019. Available from: URL https://www.R-project.org/.

41. Python Language Reference, version 3.7.4. [Internet]. Python Software Foundation.; Available from: Available at http://www.python.org

42. Rondeau V, Gonzalez JR. frailtypack: A computer program for the analysis of correlated failure time data using penalized likelihood estimation. Computer Methods and Programs in Biomedicine. 2005 Nov 1;80(2):154–164.

43. Rondeau V, Marzroui Y, Gonzalez JR. frailtypack: An R Package for the Analysis of Correlated Survival Data with Frailty Models Using Penalized Likelihood Estimation or Parametrical Estimation. Journal of Statistical Software. 2012 Apr 17;47(1):1–28.

44. Briggs J. 2020. Sex_based_differences. Github. https://github.com/EPPIcenter/sex_based_differences.git.cf6c325.

